# Digital Detection Meets Crisis Intervention: A National County-level study of GoGuardian Beacon implementation and sustainment on Youth Suicide Rates

**DOI:** 10.1101/2025.04.22.25326214

**Authors:** Xiaoyue Zhu, Karen Wood, Rachel Ahrens, Anas Belouali, Benjamin Batorsky, Holly C. Wilcox

## Abstract

Over the time of the COVID-19 pandemic, many school systems started to utilize educational software to identify students actively planning suicide and other acts of violence. This study examines associations between county-level youth suicide rates and the implementation of GoGuardian Beacon, a school-based software using machine learning methods for identifying students at risk for suicide. Using difference-in-differences and event study methods, we analyzed 2018-2022 suicide data comparing 70 counties with sustained Beacon implementation to 1,215 matched comparison counties that never implemented Beacon. In our primary analysis, counties that maintained consistent Beacon use had 24.4% lower youth suicide rates during 2021–2022 (p < 0.05). In sensitivity analyses defining implementation based on initial adoption regardless of subsequent use, the association was attenuated and not statistically significant. Taken together, these findings indicate that counties with sustained use of Beacon had lower youth suicide rates in our primary analyses, while also highlighting the possibility that broader contextual factors (e.g., local mental health infrastructure and school system characteristics) contribute to the observed differences. Randomized trials with prospective follow-up, more information on school and community resources, and quality of Beacon response pathways after identification are needed to understand the effect of Beacon and clarify the independent contribution of digital monitoring tools within comprehensive youth suicide prevention strategies.

## Introduction

In October 2021, the American Academy of Pediatrics (AAP), the American Academy of Child and Adolescent Psychiatry (AACAP), and the Children’s Hospital Association (CHA) declared a national state of emergency in children’s mental health, emphasizing the important role of schools in addressing the mental health crisis among children (American Academy of Pediatrics et al., 2021). These agencies emphasized that schools are important settings for identifying and delivering mental health services and support, as many children spend significant time in educational settings. Suicide remains a leading cause of death among young people: in 2023, it was the second leading cause of death for those aged 10-14 and 15-24 in the United States (CDC, 2025). A recent analysis of the U.S. National Violent Death Reporting System (NVDRS) found that most youth who died by suicide had no history of mental health treatment, and fewer than one-third were receiving treatment in the months prior to their death (Fontanella et al., 2025). This highlights a critical gap in our ability to identify and intervene during periods of acute suicide risk, a gap that persists largely because we lack tools capable of detecting risk and delivering resources in real time.

Digital technology, particularly when coupled with advances in machine learning and natural language processing, offers a promising avenue for continuous suicide-risk identification and timely intervention. Given that the majority of suicides occur on an individual’s first attempt (Bostwick et al., 2016), sectors serving children, such as school systems, should establish clear and robust pathways for risk identification and response. Schools are increasingly being expected to play a role in addressing youth mental health: in 2023, 2,070 school-aged children (6-18 years) died by suicide, and suicide rates among 10-18-year-olds rose steadily from 2007 through 2018, with fluctuations thereafter (CDC, 2025; Curtin et al., 2023). Moreover, in 2023, 22% of high school students reported seriously considering suicide, and 10% reported attempting suicide in the past year (CDC, 2024).

The last decade, especially during the COVID-19 pandemic, has seen a dramatic increase in student use of school-issued computing devices, spurring interest in digital tools that can identify at-risk students and connect them with support. GoGuardian Beacon (hereafter Beacon) is one such tool, designed to detect online activity on school-managed devices, including search queries, web browsing, email, and web applications that may indicate potential risk of suicide, self-harm, violence or harm to others. Schools using Beacon receive alerts regarding flagged activity to designated staff members, who can then determine appropriate follow-up actions. A recent report indicates that AI-based suicide risk monitoring can effectively identify at-risk students but research is needed on the safety, effectiveness and outcomes of these tools (Ayer et al., 2023). This study examines trends in youth suicide rates from 2018 to 2022 across U.S. counties where Beacon was implemented versus those without Beacon implementation. Using difference-in-differences approaches, we assess differences in youth suicide rates between counties that implemented and sustained Beacon versus control counties that never implemented Beacon. We performed comprehensive sensitivity analyses and analysis of pre-intervention trends to examine multiple cohort definitions and outcome specifications to evaluate the robustness of observed associations. Given the non-random nature of Beacon adoption and the challenges inherent in observational implementation research, this analysis also explores patterns of technology adoption while adjusting for county-level characteristics that may influence youth mental health outcomes. The findings are, as far as we are aware, the first to study the effect of such school-based AI tool on youth suicide, and call for future randomized studies to investigate the safety and effectiveness of such tools and the resulting workflows in preventing youth suicide.

## Materials and Methods

### Data Sources

This study utilizes restricted county-level suicide mortality data for youths aged 5-19, obtained from the Centers for Disease Control and Prevention (CDC) for all U.S. counties from 2018 to 2022. The dataset includes county Federal Information Processing System (FIPS) codes, youth suicide rates (expressed as the number of suicides per 100,000), and county-level population data.

Beacon implementation data were obtained from internal GoGuardian enterprise sources that track product usage. These data include the number of Beacon licenses purchased, the number of students actively monitored, and aggregated counts of alerts related to suicide risk. These variables were linked to the CDC dataset at the county level using FIPS codes to examine patterns in suicide rates relative to Beacon adoption.

Additional data sources were integrated for Generalized Linear Model (GLM) fitting and county-specific covariate analysis. Data from the National Center for Education Statistics (NCES, 2016-2018) include county-level demographic variables for this period (see Table 1). Additionally, counties were classified into one of eight regions: New England, Mideast, Great Lakes, Plains, Southeast, Southwest, Rocky Mountain, or Far West, based on the Bureau of Economic Analysis Regional Classifications.

**Table 1:** List of county-level demographic variables and their sources.

| **Variable** | **Years Averaged** | **Source** |
| --- | --- | --- |
| Fraction of Population of Different Races/Ethnicities | 2017-2018 | US Census Bureau |
| Population | 2017-2018 | US Census Bureau |
| Median Household Income | 2017-2018 | US Census Bureau |
| Percent of Families below Poverty Line | 2017-2018 | US Census Bureau |
| Unemployment Rate | 2017-2018 | US Census Bureau |
| Largest District Size | 2016-2018 | NCES |
| Fraction of Students on Free or Reduced Price Lunches | 2016-2018 | NCES |
| Urbanicity | 2013 | US Census Bureau |
| Region | N/A | Bureau of Economic Analysis |

### County Selection and Treatment Classification

Counties with fewer than 3,000 youths were excluded from the analysis to reduce variability in suicide rate estimates due to small sample sizes. Counties were classified as having implemented Beacon if they met predefined usage thresholds, which were established to ensure meaningful exposure to the tool. These criteria included:

1. Reaching the 5th percentile or higher in total reported alerts categorized as “awareness” (i.e., action taken on an alert) within three months of initial Beacon purchase.
2. Maintaining usage for at least 24 months, with no more than 25% of months classified as inactive. Given the nature of school years, it is expected that there will be a dormant period in which most schools are not in session.
3. Implementing Beacon between July 2020 and December 2020, ensuring that both pre-implementation and post-implementation suicide rate data were available. This period was chosen as a reasonable midpoint within the CDC data range from 2018 to 2022.

Thus, treated counties began monitoring students with Beacon between August and December 2020 and maintained usage for at least 75% of the months between January 2021 and December 2022 (n=65). Counties that did not implement Beacon before January 2023 were classified as controls. These definitions were applied to ensure that comparisons were made between counties with sustained Beacon usage and those without documented exposure to the tool (n=1,236).

### Temporal Framework and COVID-19 Context

To account for potential confounding effects of the COVID-19 pandemic, the analysis divided the study period into four timeframes:

- **Pre-pandemic baseline (February 2018–February 2020):** Provides a reference period before COVID-19 disruptions.
- **Peak pandemic (March 2020–July 2020):** Captures the initial phase of widespread uncertainty and social disruption.
- **Back-to-school transition (August 2020–December 2020):** Reflects the period when many schools resumed in-person or hybrid learning.
- **Post-pandemic adjustment (January 2021–December 2022):** Represents the longer-term period following the pandemic’s peak effects.

This segmentation allows for an examination of whether trends in suicide rates differed between counties that implemented Beacon and those that did not while accounting for broader societal changes.

### Statistical Analyses

Monthly youth suicide rates between treatment and control counties in different timeframes were first examined using two-sample independent t-tests (with SciPy). A one-sided alternative hypothesis was specified. We assessed the equality of variances using Levene’s test at a significance level of 0.05. For timeframes where Levene’s test produced a p-value greater than 0.05, we assumed equal variances and applied the standard two-sample t-test. Conversely, if the p-value was less than 0.05, indicating unequal variances, we used Welch’s t-test.

To account for county-level demographic and regional differences between counties, a Poisson GLM was used within a difference-in-differences framework to estimate associations between Beacon implementation and youth suicide rates. We first modeled the suicide rate per 100,000 youths directly:

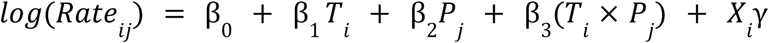

Where,

- *i* indexes counties and *j* indexes timeframes (pre-intervention: February 2018 - February 2020; post-intervention: January 2021 - December 2022)
- *T*_*i*_: Binary treatment indicator (1 if county *i* implemented Beacon meeting the *i* criteria described above, 0 otherwise)
- *P*_*j*_: Binary post-intervention indicator (1 for January 2021 - December 2022, 0 for *j* February 2018 - February 2020)
- β_3_: Difference-in-differences coefficient, representing the treatment effect
- *X*_*i*_: Vector of county-level demographic and socioeconomic covariates measured during the pre-intervention period, including racial and ethnic composition, population size, median household income, poverty rate, and unemployment rate (see Table 2 for a complete list)

**Table 2:** Comparison of average youth suicide rates (per 100,000 per month per county) between control and treatment groups across four timeframes. Values represent mean ± standard error of the mean (SEM). P-values from one-sided independent t-tests (treatment < control). Significance: *p < 0.05,**p < 0.01, ***p < 0.001.

| Timeframe | Control<br>*n* = 1,215 | Treatment<br>*n* = 70 | p-value |
| --- | --- | --- | --- |
| Pre-pandemic baseline (February 2018 - February 2020) | 0.504 ± 0.019 | 0.525 ± 0.051 | 0.645 |
| Peak pandemic (March 2020 - July 2020) | 0.457 ± 0.037 | 0.347 ± 0.081 | 0.237 |
| Back-to-school transition (August 2020 - December 2020) | 0.485 ± 0.035 | 0.458 ± 0.121 | 0.425 |
| Post-pandemic adjustment (January 2021 - December 2022) | 0.482 ± 0.017 | 0.364 ± 0.039 | < 0.01** |

Standard errors were clustered by county to account for within-county correlation across timeframes, with inference conducted using t-distributions with degrees of freedom equal to the number of counties minus one.

### Sensitivity Analyses

To assess the robustness of our findings, we conducted a comprehensive series of sensitivity analyses examining multiple analytic approaches, outcome specifications, and statistical inference methods.

#### Cohort Definitions

We compared two distinct approaches to defining treatment and control groups:

- **Per-Protocol (PP) Analysis.** Our primary analysis employed a strict per-protocol design requiring counties to meet predetermined implementation fidelity criteria as previously described. It yielded 70 treatment counties and 1,212 control counties.
- **Intent-to-Treat (ITT) Analysis**. It classified all counties with any documented Beacon exposure during the study period as treated (n=154), regardless of implementation fidelity or sustained usage. This more inclusive definition assessed the effect of Beacon availability rather than sustained implementation.

#### Outcome Specifications

We examined two specifications of the primary outcome:

1. **Suicide Rate.** Computed monthly youth suicide rates per 100,000 population at the county level.
2. **Count with Offset**. To properly account for the discrete nature of suicide counts and varying county exposure times, we employed a count-based approach using Poisson regression with an offset term:

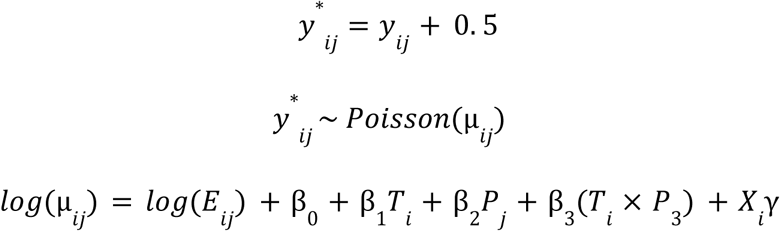 where *y_*i*j_* is the observed youth suicide count in county *i* during timeframe

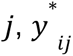 is the continuity-corrected count, µ*_ij_* is the expected count, and *E*_*ij*_ = *population*_*ij*_ × *months*_*j*_ represents person-months at risk.

### Primary Analysis

To examine the temporal dynamics of treatment effects and assess the parallel trends assumption, we conducted an event study using the per-protocol cohort and count-with-offset method across all four study timeframes. We estimated a Poisson GLM with timeframe-specific treatment effects:

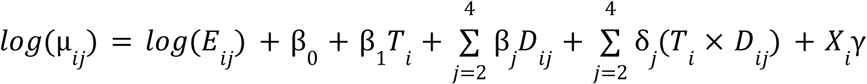

where *T*_*i*_ is the treatment indicator, *D_*ij*_* are timeframe indicators (omitting the baseline period), and *X*_*i*_ includes pre-intervention county-level covariates. The coefficients δ_*j*_ represent timeframe-specific treatment effects relative to baseline, with rate ratios computed as *RR*_*j*_ = *exp*(δ_*j*_). Parallel trends were assessed by examining δ_2_ and δ_3_ (Spring / Summer 2020 and Fall 2020), when most treatment counties had not yet fully implemented Beacon. Significant coefficients during these periods would indicate pre-existing differential trends between treatment and control counties. Standard errors were estimated using two-way clustering by county and timeframe, with t-based inference using conservative degrees of freedom *min*(*G*_1_− 1, *G*_2_− 1), where *G*_1_ and *G*_2_ are the number of county and timeframe clusters, respectively.

## Results

To account for potential confounding effects of the COVID-19 pandemic and align with Beacon implementation patterns, we segmented the study period into four distinct timeframes. Table 2 presents mean monthly suicide rates across these four timeframes. In the pre-pandemic baseline period (February 2018 - February 2020), suicide rates were nearly identical between treatment and control counties (treatment = 0.525 ± 0.051, control = 0.504 ± 0.019, p=0.645, one-sided t-test). However, during the post-pandemic adjustment period (January 2021 - December 2022), treatment counties exhibited substantially lower rates (treatment = 0.364 ± 0.039, control = 0.482 ± 0.017, p=0.003), representing a 24.4% relative difference (Figure 1).

**Figure 1:**
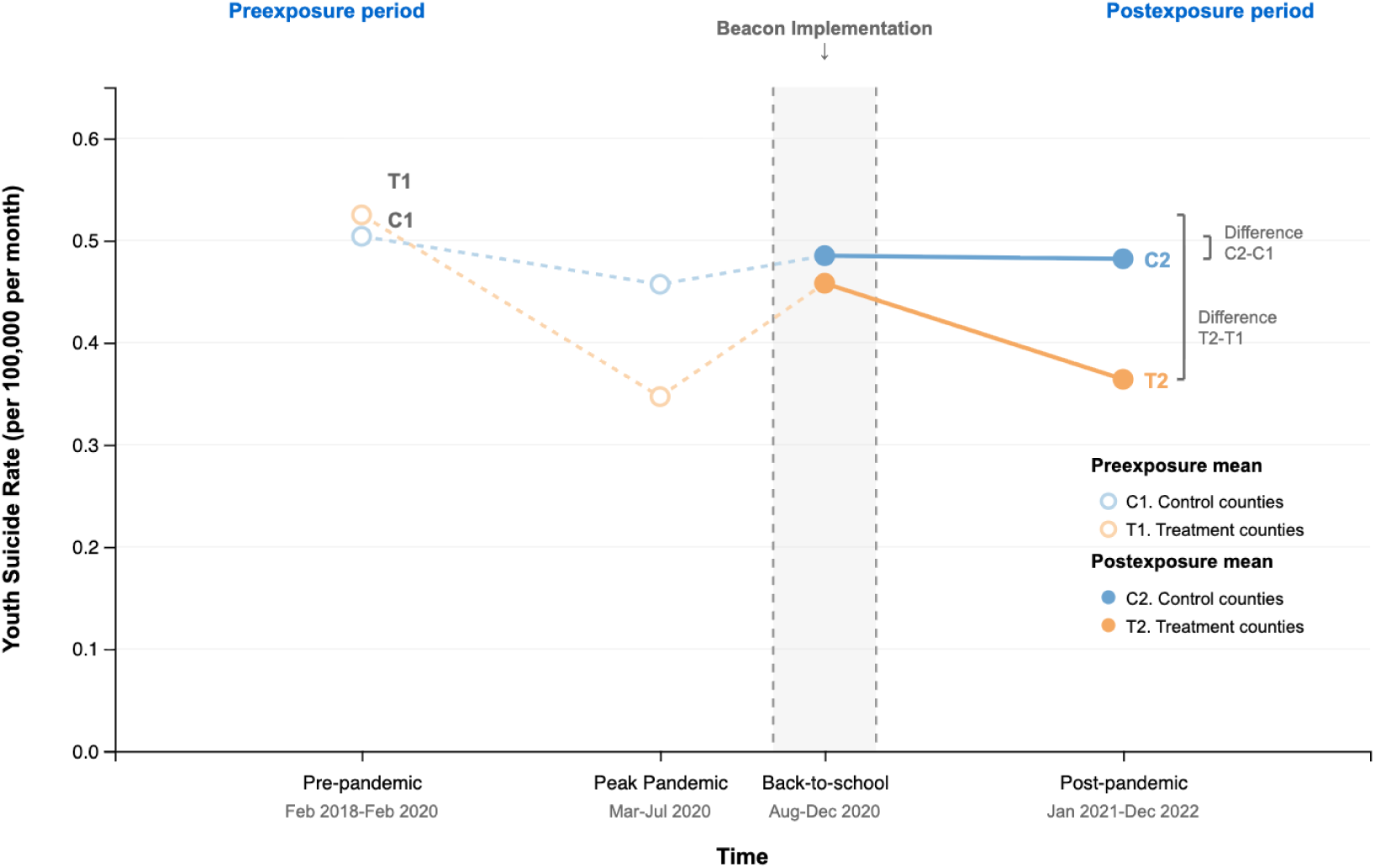
Beacon Implementation and Youth Suicide Rate. The difference-in-differences pattern comparing youth suicide rates between treatment counties and control counties. Data points represent mean monthly suicide rates per 100,000 youth aged 5-19, with C1 & T1 indicating pre-exposure baselines and C2 & T2 indicating post-exposure outcomes. The Beacon implementation period (Jul - Dec 2020) is highlighted in a gray shaded band. Vertical brackets display within-group changes over time (C2-C1 for control counties; T2-T1 for treatment counties). The x-axis is scaled proportionally to reflect actual time durations in months across the study period (2018 - 2022).

Using a difference-in-differences Poisson GLM that adjusts for county-level demographic and regional characteristics, we found that counties implementing Beacon exhibited lower suicide rates in the post-pandemic adjustment period relative to control counties. The interaction term between Beacon implementation and the post-intervention timeframe was statistically significant in the per-protocol rate model (RR = 0.725 (0.560, 0.938), p<0.05; Table 3 first row).

**Table 3:** Sensitivity analysis of the difference-in-differences treatment effect across cohort definitions and outcome specifications. All models include county-level demographic covariates and county-clustered standard errors. Rate models use suicide rates per 100,000; Count + offset models use death counts with log(population × months) offset. RR = rate ratio for the treatment × post-period interaction term. T = treatment group; C = control group.

| Approach | Model | Sample | RR (95% CI) | p-value |
| --- | --- | --- | --- | --- |
| Per-Protocol | Rate | 70 T / 1,215 C | 0.725 (0.560, 0.938) | < 0.05* |
|  | Count + offset |  | 1.041 (0.891, 1.215) | 0.616 |
| Intent-to-Treat | Rate | 836 T / 1,215 C | 0.992 (0.869, 1.133) | 0.907 |
|  | Count + offset |  | 1.028 (0.963, 1.098) | 0.408 |

To assess the robustness of this finding, we conducted a comprehensive sensitivity analysis (Table 3). The results varied substantially between the model specifications and the definitions of the cohorts. The per-protocol analysis × suicide rate model combination yielded a statistically significant difference-in-differences effect, as reported previously (p<0.05). However, this effect was not replicated when using the count-with-offset specification, which is more widely adopted for rare counts data (RR = 1.041 (0.891, 1.215), p=0.616).

The per-protocol approach assigns treatment based on post-implementation behaviors (e.g., sustained Beacon usage), which may introduce selection bias that inflates the treatment effect. An intent-to-treat approach treats any county implementing Beacon during July - December 2020 as “treated” regardless of subsequent usage patterns. This design reduces bias from conditioning on post-treatment variables. The intention-to-treat approach showed no significant association in either the rate model (RR = 0.992 (0.869, 1.133), p=0.907) or the count-with-offset model (RR = 1.028 (0.963, 1.098), p=0.408). The divergence between rate and count-based specifications, as well as between per-protocol and intent-to-treat approaches, suggests that the initial significant effect of Beacon is sensitive to both outcome measurement and cohort definition.

Further event study analysis revealed evidence of pre-existing differences between treatment and control counties that preceded Beacon implementation (Table 4). During the peak pandemic period (March 2020 - July 2020), when most treatment counties had not yet achieved full implementation, treated counties already exhibited significantly lower suicide rates relative to the baseline period (RR = 0.598 (0.478–0.749, p<0.01). This pattern persisted during the back-to-school transition period (August 2020 - December 2020), where most active implementation happened (RR = 0.660 (0.519–0.840), p<0.05). In the post-pandemic adjustment period (January 2021 - December 2022), the treatment effect was no longer statistically significant (RR = 1.039 (0.851–1.269), p=0.585). These results indicate a violation of the parallel trends assumption underlying the difference-in-differences design, that the treatment counties were on a different trajectory before full Beacon implementation. Overall, it suggests that the observed associations reflect pre-existing differences rather than a causal effect of the full Beacon implementation.

**Table 4:** Event study analysis of timeframe-specific treatment effects relative to baseline period. Rate ratios (RR) represent the multiplicative change in suicide rates for treatment counties relative to control counties in each timeframe compared to the baseline period (February 2018 - February 2020). All models include county-level demographic covariates. Per-protocol analysis used two-way clustering (county by timeframe, df = 3); intent-to-treat analysis used county clustering (df = 2, 047). T = treated counties; C = control counties.

| Approach | Timeframe | RR (95% CI) | p-value |
| --- | --- | --- | --- |
| *Baseline: February 2018 – February 2020 (reference)* |  |  |  |
| Per-Protocol<br>(70 T / 1, 215 C) | March - July 2020 | 0.598 (0.478, 0.749) | < 0.01** |
|  | August - December 2020 | 0.660 (0.519, 0.840) | < 0.05* |
|  | January 2021 - December 2022 | 1.039 (0.851, 1.269) | 0.585 |
| Intent-to-Treat<br>(836 T / 1, 215 C) | March - July 2020 | 0.662 (0.608, 0.721) | < 0.001*** |
|  | August - December 2020 | 0.747 (0.689, 0.810) | < 0.001*** |
|  | January 2021 - December 2022 | 1.028 (0.962, 1.098) | 0.418 |

## Discussion

This study examined associations between county-level youth suicide rates and the implementation of GoGuardian Beacon, a digital suicide risk monitoring tool, across U.S. counties from 2018 to 2022. While suicide rates increased nationally in 2021 (Stone, 2023), counties that had implemented GoGuardian Beacon with high fidelity exhibited lower youth suicide rates during the post-pandemic period compared to those that had not. Comprehensive sensitivity analyses and event study designs revealed that these associations are not robust to different model specifications and appear to reflect pre-existing differences rather than intervention effects.

Our results indicate that counties implementing Beacon with sustained usage had lower youth suicide rates during 2021 - 2022 compared to counties without. However, event study analyses showed that these counties exhibited lower rates even during the preceding summer and fall. This temporal pattern suggests that observed differences may reflect pre-existing county characteristics rather than intervention effects. For example, school districts that make the decision to utilize Beacon may have experienced student suicides or have had more mental-health-related emergencies. They may also have more resources for addressing student mental health challenges.

The impact of the identification of students using AI methods on suicide rate depends on the referral pathway at school. Communities throughout the United States have various access to resources and mental health services. Beacon adoption may serve as a marker for counties with stronger mental health infrastructure. Such counties may possess dedicated mental health personnel, established response protocols, stronger school-community partnerships, and more comprehensive prevention frameworks. These same characteristics that enable successful Beacon implementation may independently reduce youth suicide rates through multiple parallel mechanisms. During the study period (2018 - 2022), many counties implemented concurrent mental health initiatives, expanded school counseling services, or strengthened mental health partnerships, all confounding variables that our data cannot observe or disentangle from Beacon’s contributions.

Further evidence for this interpretation comes from the sensitivity analyses. The initial significant association was limited to a specific combination of modeling choices: per-protocol definition with suicide rate as the outcome. When using count-based models with count offset to model for rare count events, or when using an intent-to-treat approach that avoids conditioning on post-implementation behaviors, no significant associations were found. Thus, the observed associations are concentrated exclusively among counties achieving sustained implementation, exactly the pattern expected when unmeasured confounding variables likely drive both implementation fidelity and outcomes.

Schools are increasingly taking a proactive role in identifying students experiencing distress or risk for self-directed or other-directed violence. Schools frequently employ multiple, concurrent approaches for mental health and suicide prevention, often initiated or terminated across different periods based on leadership priorities and resources. This complexity makes it challenging to disentangle the specific effects of any single program from the influence of other interventions and contextual factors on youth suicide rates. Suicide prevention advocacy organizations consistently advise schools to utilize a coordinated prevention approach that integrates multiple elements, such as teacher training, direct student interventions, embedded mental health providers, and robust partnerships with community-level mental health organizations (U.S. Department of Health and Human Services, 2024). Consequently, isolating the impact of any singular school-based approach from this web of elements is inherently difficult. Even a well-designed randomized trial, which might attempt to match and randomly assign schools or districts, would still face significant threats to validity from various influential contextual variables. This paper, therefore, represents one of the initial evaluations of the impact of this widely utilized, coordinated prevention framework in the United States. Factors such as access to mental healthcare, availability of firearms and other lethal means, as well as broader economic and family-level influences, are critical county-level variables that may also interact with and affect youth suicide rates (Steelesmith et al., 2019).

Evaluating the impact of interventions on rate outcomes, such as youth suicide, is inherently challenging. The national suicide statistics at the county level are suppressed when counts are small. As such, we used suicide rates and focused on counties with greater student representation. This approach excluded rural counties where suicide rates are higher and likely underestimates the true impact of Beacon due to the mismatch between the intervention unit and the data aggregation unit. Specifically, some counties were treated as “Beacon counties” even if the intervention is utilized by a district covering only one-third of the county. Additionally, schools have varying workflows related to Beacon. Some schools paid for an enhanced 24/7 service to facilitate the referral workflow, while others relied solely on embedded personnel to manage and address referrals. Similarly, some school districts maintain Beacon as a 24/7 service, whereas others restrict its operation to school hours, suspending its use during evenings, holiday breaks, or summer recess. In prevention science, the impact on outcomes rests on the quality of implementation. Except for Beacon usage patterns over the study period, details on the implementation quality of the counties are lacking. Schools in control counties could have been using other similar software (e.g., Gaggle, Bark), but we did not have data on the utilization of these tools to adjust for this.

From a policy perspective, these findings have important implications. First, the absence of causal evidence for Beacon’s effectiveness does not imply that digital monitoring tools lack value. Rather, it reflects the limitations of observational designs for isolating their specific contributions within complex prevention ecosystems. Counties considering Beacon adoption should view it as a part of comprehensive suicide prevention strategies, not a standalone solution. Second, it suggests that implementation support and school resources may be as critical as the technology itself. Counties lacking mental health infrastructure and experienced staff may struggle to implement Beacon effectively. Third, our findings highlight the need for more rigorous evaluation designs. Randomized controlled trials or regression discontinuity approaches leveraging natural thresholds in adoption decisions will provide better evidence for the intervention effect. Such designs are particularly important given the high stakes of suicide prevention and the substantial investments counties make in digital monitoring systems.

As one of the first large-scale, quantitative analyses examining digital suicide monitoring in school settings, this study serves as a foundation for future research. First, the current analysis relied on county-level suicide data, which neglects heterogeneity in school-specific implementation practices and student-level outcomes. Future research should examine implementation fidelity at the school and district levels, response protocols following alerts, and pathways from detection to intervention. Second, while we controlled for demographic and socioeconomic characteristics, unmeasured confounding remains. County-level access to lethal means, mental health policies and resources and prevention programming could not be quantified from available data. Longitudinal studies with more granular measures of such variables could better isolate the contribution of specific components. Finally, the COVID-19 pandemic introduced significant disruptions to schools and mental health infrastructure, complicating the interpretation of trends during our study period. While we segmented the analysis into pandemic-relevant timeframes and included them in event study models, residual confounding factors cannot be ruled out. Future evaluations conducted during stable periods may yield different patterns. Furthermore, this type of monitoring intervention was highly relevant at the time of the pandemic, amidst a youth mental health crisis, and shift to online learning. However, the direct applicability of Beacon implementation post-pandemic is another consideration. Partnerships are needed between industry and academic researchers in the suicide prevention and public health space. For this independent evaluation, GoGuardian was willing to share their proprietary data and demonstrated a commitment to transparency and safety. This study can be considered a quality improvement and quality control project to understand the impact of GoGuardian Beacon on student outcomes.

## Data Availability

The data used in this study include restricted CDC datasets and proprietary product data, and are not publicly available. Due to privacy and data use limitations, the data cannot be shared.

## Compliance with Ethical Standards

### Funding

GoGuardian provided time for employees X.Z., R.A., K.W., and B.B. to work on this paper as part of their quality improvement mission. A.B.’s contribution to this study was supported by NLM doctoral training grant T15LM013979. H.C.W.’s contributions to this study were in kind.

GoGuardian provided proprietary data on Beacon usage to Johns Hopkins authors to conduct replication and independent analyses.

### Ethics approval

This study was approved by the Centers for Disease Prevention and Control.

### Conflicts of interest / Competing interests

X.Z and B.B are current employees of GoGuardian. K.W. and R.A. are previous employees of the company. Johns Hopkins University collaborators, A.B. and H.C.W., received no external funding or support from GoGuardian or, for H.C.W., other agencies for her contributions to this study.

A.B. independently conducted analyses using company-provided, de-identified aggregates and confirmed the reported results; all analytic decisions and interpretations in the manuscript were made jointly by the author team.

### Consent to participate

Not Applicable

## References

1. American Academy of Pediatrics, American Academy of Child and Adolescent Psychiatry, & Children’s Hospital Association. (2021). AAP-AACAP-CHA declaration of a national emergency in child and adolescent mental health. https://www.aap.org/en/advocacy/child-and-adolescent-healthy-mental-development/aap-aacap-cha-declaration-of-a-national-emergency-in-child-and-adolescent-mental-health/

2. Centers for Disease Control and Prevention, National Center for Injury Prevention and Control. (2025). Web-based Injury Statistics Query and Reporting System (WISQARS). U.S. Department of Health and Human Services. Retrieved December 13, 2025, from https://wisqars.cdc.gov/

3. Fontanella, C. A., Xia, X., Campo, J. V., Steelesmith, D. L., Bridge, J. A., & Ruch, D. A. (2025). Characteristics associated with mental health treatment prior to suicide among youth in the United States. Journal of the American Academy of Child & Adolescent Psychiatry, 64(5), 625–635. 10.1016/j.jaac.2024.07.921

4. Bostwick, J. M., Pabbati, C., Geske, J. R., & McKean, A. J. (2016). Suicide attempt as a risk factor for completed suicide: even more lethal than we knew. American journal of psychiatry, 173(11), 1094–1100. 10.1176/appi.ajp.2016.15070854

5. Curtin, S. C., Garnett, M. F., & Ahmad, F. B. (2023). Provisional estimates of suicide by demographic characteristics: United States, 2022 (Vital Statistics Rapid Release No. 34). National Center for Health Statistics. 10.15620/cdc:133702

6. Centers for Disease Control and Prevention. (2024). Youth Risk Behavior Survey Data Summary & Trends Report: 2013-2023. https://www.cdc.gov/yrbs

7. Ayer, L., Boudreaux, B., Paige, J. W., Holmes, P., Blagg, T. L., & Mendon-Plasek, S. J. (2024). Artificial Intelligence–Based Student Activity Monitoring for Suicide Risk: Considerations for K–12 Schools, Caregivers, Government, and Technology Developers. Rand health quarterly, 11(2), 2. https://pmc.ncbi.nlm.nih.gov/articles/PMC10911757/

8. Stone, D. M. (2023). Notes from the field: recent changes in suicide rates, by race and ethnicity and age group—United States, 2021. MMWR. Morbidity and Mortality Weekly Report, 72. 10.15585/mmwr.mm7206a4

9. U.S. Department of Health and Human Services. (2024). National strategy for suicide prevention. https://www.hhs.gov/programs/prevention-and-wellness/mental-health-substance-use-disorder/national-strategy-suicide-prevention/index.html

10. Steelesmith, D. L., Fontanella, C. A., Campo, J. V., Bridge, J. A., Warren, K. L., & Root, E. D. (2019). Contextual factors associated with county-level suicide rates in the United States, 1999 to 2016. JAMA network open, 2(9), e1910936–e1910936. 10.1001/jamanetworkopen.2019.10936

